# Interest of structured reporting and combined automated co-registration and lesion color-coding maps for longitudinal magnetic resonance imaging analysis in patients with multiple sclerosis : the MS-LOBI-SR study protocol

**DOI:** 10.1101/2022.07.02.22276009

**Authors:** Sébastien Kerdraon, Ichem Mohtarif, Kevin Rose, Douraïed Ben Salem, Brieg Dissaux, Julien Ognard

## Abstract

MS is characterized by chronic and immune-mediated inflammatory lesions of the brain white matter, disseminated in time and space. As the technology improved over the next three decades, MRI quickly grew to become the single most important paraclinical diagnostic and monitoring tool available. A lesion will be defined as having a high T2/FLAIR signal ≥ 3 millimeters in diameter. The purpose of this study, witch includes 95 patients, was to investigate the benefit of a computer assisted detection reading using combined automated co-registration and lesion color-coding method approach, over a conventional reading approach in follow-up examinations of patients with MS, especially regarding diagnostic accuracy, inter/intra-reader agreement and required reading time. The impact of the use of a structured report on these variables will also be evaluated. The main judgment criteria will be : diagnostic accuracy (effectiveness) to assess the progression-activity of the disease based on the appearance of a T2w-FLAIR hyperintensity or a T1w enhancement on the follow-up examination, compared between consensus corrected CADR and CR readings.The reading of the examinations will be made by radiologists and radiology technician.

## Introduction

MS is characterized by chronic and immune-mediated inflammatory lesions of the brain white matter (WM), disseminated in time and space (1). Besides cognitive impact, MS leads to the highest number of permanent disability in young adults (2). Europe rank among the regions with its highest incidence and prevalence (3).

The diagnosis is based on a bundle of clinical, paraclinical and evolutionary arguments. As the technology improved over the next three decades, MRI quickly grew to become the single most important paraclinical diagnostic and monitoring tool available. The ability to demonstrate lesion dissemination in space and time has led to the common use of MRI as a paraclinical measure to support a diagnosis of MS, including in patients with clinically isolated demyelinating syndromes(4).

In addition, MRI is used to monitor the progress of disease in patients with clinically definite MS, including assessment of lesions and atrophy (5).

Current imaging markers with relevance for MS are associated with disease activity and progression, and include, among other features, number or volume of hyperintense brain lesions visible on T2-weighted MRI images, contrast-enhancing T1 lesions, increased annual brain volume loss and T1-hypointense “black holes” (6). All of these points position the MRI as a routine examination, and exploration of longitudinal changes in patients with MS is therefore very frequent.

Mainly the increase in the spatial resolution of acquisitions combined with the number/load of lesion may impair diagnostic accuracy due to a laborious interpretation. This visual MRI analysis is known as a tedious and time-consuming task that may suffer from a lack of sensitivity and intra- and inter-observer reproducibility. The heterogeneity of used machines (manufacturer, magnetic field) for the longitudinal follow up, also add to the variability of interpretation and can even have an impact regarding the experience of the reader. Hagens et al. (7) calculated an inter-rater agreement on involvement per anatomical region and dissemination in space and time that was moderate to good for 1.5 and 3T magnetic fields, while 3T slightly improved agreement between experienced raters, but slightly decreased agreement between less-experienced raters.

The processing of medical images makes it possible to assist in the interpretation and analysis of the images and more particularly in the longitudinal follow-up of a patient (8). This helps to smooth out differences in interpretation between readers, promotes the same interpretation at different levels of experience, and allows for an increase in reading speed. Wang et al. (9) have showed that a semi-automated assistive software that aids in the identification of new lesions has a beneficial effect for both neuroradiologists and non-neuroradiologists, though the effect is more profound in the non-neuroradiologist group.

Zopfs et al. (10) used automated co-registration and lesion color-coding of MS-associated FLAIR-lesions in followup MRI of MS patients and found that it increased diagnostic accuracy and reduced the reading time significantly. They also summed up different approaches and tools available to facilitate detection of disease progression, such as subtraction or automated co-registration techniques (Galetto AJNR 2018), voxel-based registration of pairs of images (12), subtraction of previously coregistered and intensity-normalized pairs of images (13).

With the recent advent of artificial intelligence, several machine learning techniques have been proposed to improve automated delineation of pathologies (14), however, many of these techniques may not be suitable in a clinical environment, as they either require special informatic skills to be implemented or difficult to include in a clinical workflow (15).

In recent years, structured reporting (SR) has been used in different fields of radiology to overcome report variability and increase standardization (16). Moreover, evidence is increasing that the implementation of SR may increase the standardization of reports, improve communication between the radiologist and the referring physician, potentially alter patient management, and simplify clinical decision making (17). SR may be useful in analyzing patient outcomes, which is relevant in the setting of clinical trials (18). In the context of a pathology leading to potentially high inter-observer variability, it seemed necessary to standardize the reading, using a structured report.

Therefore, the purpose of this study was to investigate the benefit of a computer assited detection reading (CADR) using combined automated co-registration and lesion color-coding method approach, over a conventional reading approach (CR) in follow-up examinations of patients with MS, especially regarding diagnostic accuracy, inter/intra-reader agreement and required reading time. The impact of the use of a structured report on these variables will also be evaluated.

## Material and methods

### Ethics

The study will be conducted in accordance with the ethical regulations of the Declaration of Helsinki including its later amendments. Considering the retrospective design, all cerebral MRI exams were already performed for clinical indications, no MRI will be conducted solely for the purpose of this study. An request of reviewing of this study plan was deposited to the CERIM (Comité d’éthique de recherche en imagerie médicale). The need of informed patient consent will be adapted to this reviewing board decision.

### Subjects

Identification of potential study subjects will be conducted based on a combined exploration of the picture archiving and communication system (PACS, Centricity, GE), the radiological information system (RIS, Xplore), and the hospital information system (MEVA) screened, for a period of time adapted to the number needed of subjects, for patients 18 years of age or older who were diagnosed with MS, that underwent two consecutive MR imaging examinations of the brain for follow-up with complete standardized protocol, already included in the OFSEP (Office Français de la Sclérose en Plaque) registry,. Exclusion criterion will be retained: examination of lesser quality not allowing a correct analysis of the images. The examinations will therefore only be the ones performed in our institution (mutli-machine, single center).

### Imaging data

Detailed scan parameters are given for the two MRI systems of the unit : Philips Elition X 3T and GE Optima MR450W 1.5T. Protocols of MRI examinations are implemented in accordance to national guidelines (OFSEP) and described in the tables below :

**Table 1.** MRI acquisition parameters

|  | DWI | T1w TFE | T2w<br>FLAIR | T1w TSE FS |
| --- | --- | --- | --- | --- |
| **1.5T** |  |  |  |  |
| **Acquisition plane** | Axial | Sagittal | Sagittal | Axial |
| **Encoding direction** | AP | AP | AP | RL |
| **FOV** | 240x265 | 240x240 | 250x250 | 256x256 |
| **Pixel size** | 1.6x2.3x<br>3 | 1x1x1 | 1x1x1 | 1x1x1 |
| **Acceleration** | Sense 2 | Cs Sense 3 | Cs Sense 7 | Cs sense 6 |
| **TE** | 66 | 3.8 | 288 | 28 |
| **TR** | 3771 | 8.3 | 8000 | 600 |
| **TI** | NA | NA | 2300 | NA |
| **NSA** | 1 | 1 | 1 | 1 |
| **Fat Suppresion** | SPIR | No | SPIR | SPIR |
| **Time** | 1'31 | 3'32 | 5'20 | 3'58 |
| **3T** |  |  |  |  |
| **Acquisition plane** | Axial | Axial | Sagittal | Sagittal |
| **Encoding direction** | AP | AP | AP | RL |
| **FOV** | 240x240 | 256x205 | 256x216 | 256x230 |
| **Pixel size** | 1,9x1,7x<br>4 | 1x1x1 | 1,2x1,2x1,6 | 1x1x1 |
| **Acceleration** | ASSET<br>2 | ASSET 1,5 | ARC 2/<br>Hypersense<br>1,1 | ARC 2 Hypersense 1,15 |
| **TE** | 69,7 | 2,9 | 136 | 15 |
| **TR** | 8500 | 7,2 | 8000 | 500 |
| **TI** |  |  | 2112 |  |
| **NSA** | 1 | 1 | 1 | 1 |
| **Fat Suppresion** |  | No | FatSat | FatSat |
| **Time** | 1'51 | 2'26 | 4'12 | 3'48 |

### Reading process

A lesion will be defined as having a high T2/FLAIR signal ≥ 3 millimeters in diameter. The presence of a new T2w-FLAIR hyperintensity lesion will be defined as nonartifactual, new, bright areas clearly visible against the background.

All scans will be centrally collected and checked for completeness. The scans will be rated independently by four raters during two central reading sessions: one experienced rater (J.O.., neuroradiologist for 5 years), two MS researchers considered as less-experienced raters (K.R., I.M., radiology residents for 3 years), and a non-radiologist (S.K., radiology technician for 10 years).

The disagreements will be resolved in consensus by two experienced raters non-implicated in the central reading, in the same reading conditions of readings (D.B.S, neuroradiologist for 20 years, B.D., neuroradiologist for 3 years).

The order and type of readings will be randomized in and between sessions, and won’t be the same for all the raters, that will be fully blinded of clinical data.

There will be four types of readings, according to the design of the study : CADR with SR, CR with SR, CADR with NSR and CR with NSR.

Readings will be performed at the same workstation under standardized reading conditions with a time interval of 12 weeks between the conventional reading and the CADR reading, to avoid a recall bias.

For radiologists, the process will consist in performing the four types of readings. The radiology technician not carrying out in his current practice the radiological reports, he will perform only the reading using SR (CADR with SR, CR with SR).

The average time required for loading the images within the PACS will be recorded in the conventional reading session. The average time required for loading the applications and processing the images will be recorded. Furthermore, the time from being presented with the images to making the diagnosis will be recorded for each follow-up examination pair. All time measurements will be performed by an assistant not involved in the readouts.

### Computer assisted detection reading CADR

To perform CADR, the readers will use a CE-certified and FDA-approved software that facilitates follow-up examinations of the same patient (MR Longitudinal Brain Imaging [LoBI]; Philips Healthcare). The software is integrated into the vendor’s image viewer (IntellisSpace Portal, Version 11; Philips Healthcare), however is not constrained to MR images generated by Philips Healthcare scanners and generally applicable. The software automatically performs a rigid co-registration. Then, the application performs an intensity normalization and subsequent subtraction of the selected sequences. After co-registration, normalization, and subtraction, both sequences are linked at the same anatomic level, allowing manual correction if necessary. Additionally, the software creates an overlay map, which highlights a focal increase in signal intensity in red and a focal decrease in signal intensity in blue, respectively.The color intensity of the color-coding can be adjusted seamlessly. Both linked sequences and the overlay map are displayed side by side and can be viewed simultaneously on the same anatomic level. (19)

**Fig 1.**
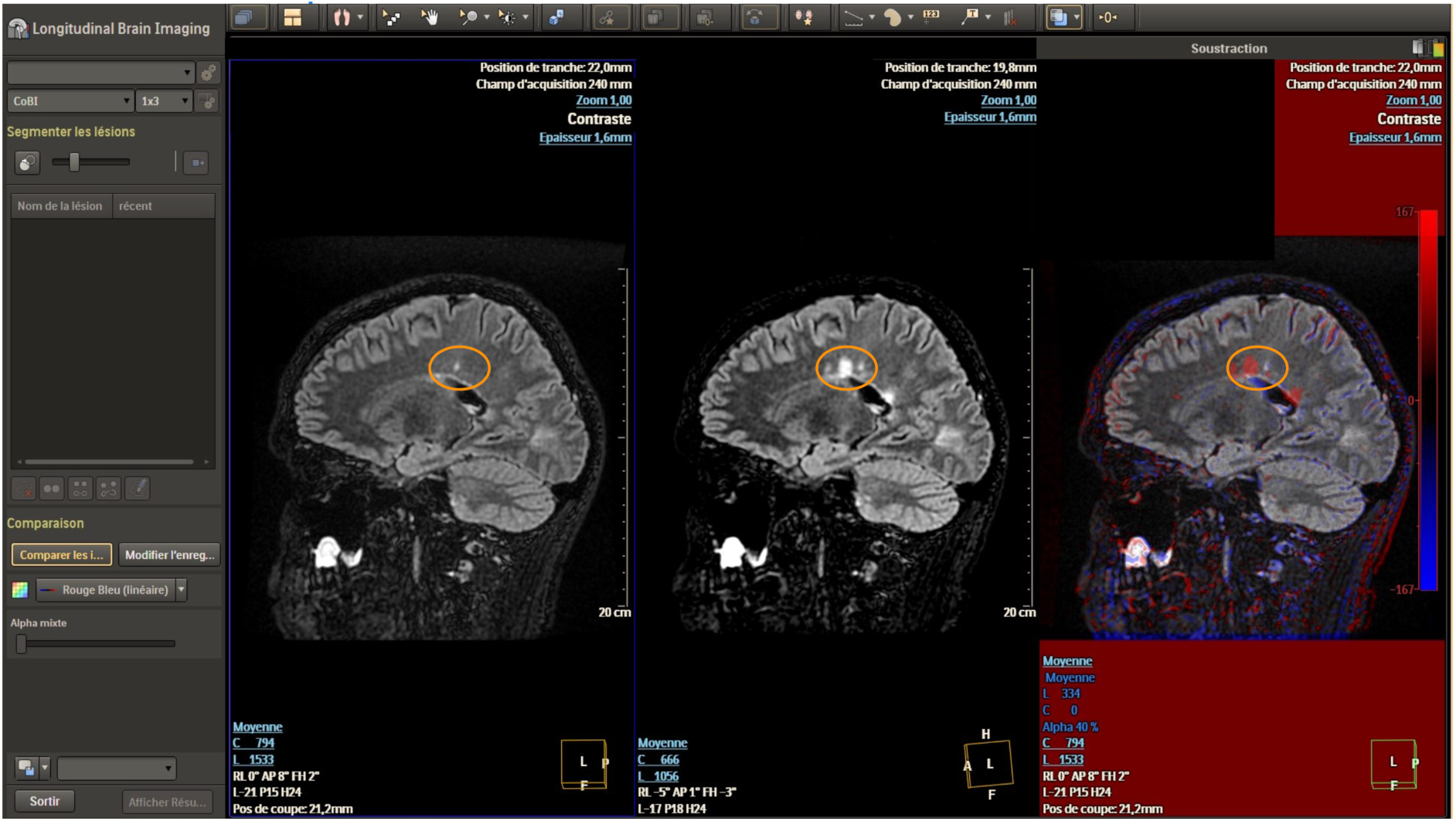
LOBI Interface. The yellow circle shows the apparition of a new hypersignal.

### Conventional reading CR

Conventional readings will be performed as the reader habits, using CUV software (Centricity Universal Viewer, GE Healthcare)

### Structured report SR

The SR will be generated using RADR software, a web application (https://radr.fr/, accessed on the 2022-06-05) that allows via a simple interface to creates reports through point and click. For the MS module, the anonymization, clinical information (such as type of MS, treatment, recent attack or not, EDSS score) as well as the sequences carried out are informed. The interface permits to select via simple clicks pre parcellated areas of the brain (lobes, corpus callosum, brainstem), and category of WM or cortex involved. The user can also inform the apparition of the lesion. A structured report is generated. A standardized table can be exported as a .Json file. For all scans, the number of inflammatory lesions larger than 3 mm in size will be scored and categorized according to the anatomical region. The number of enhancing lesions per region will be also reported if contrast media was used during the examination. Activity-progression of the disease will then be determined by each reader.

**Fig2.**
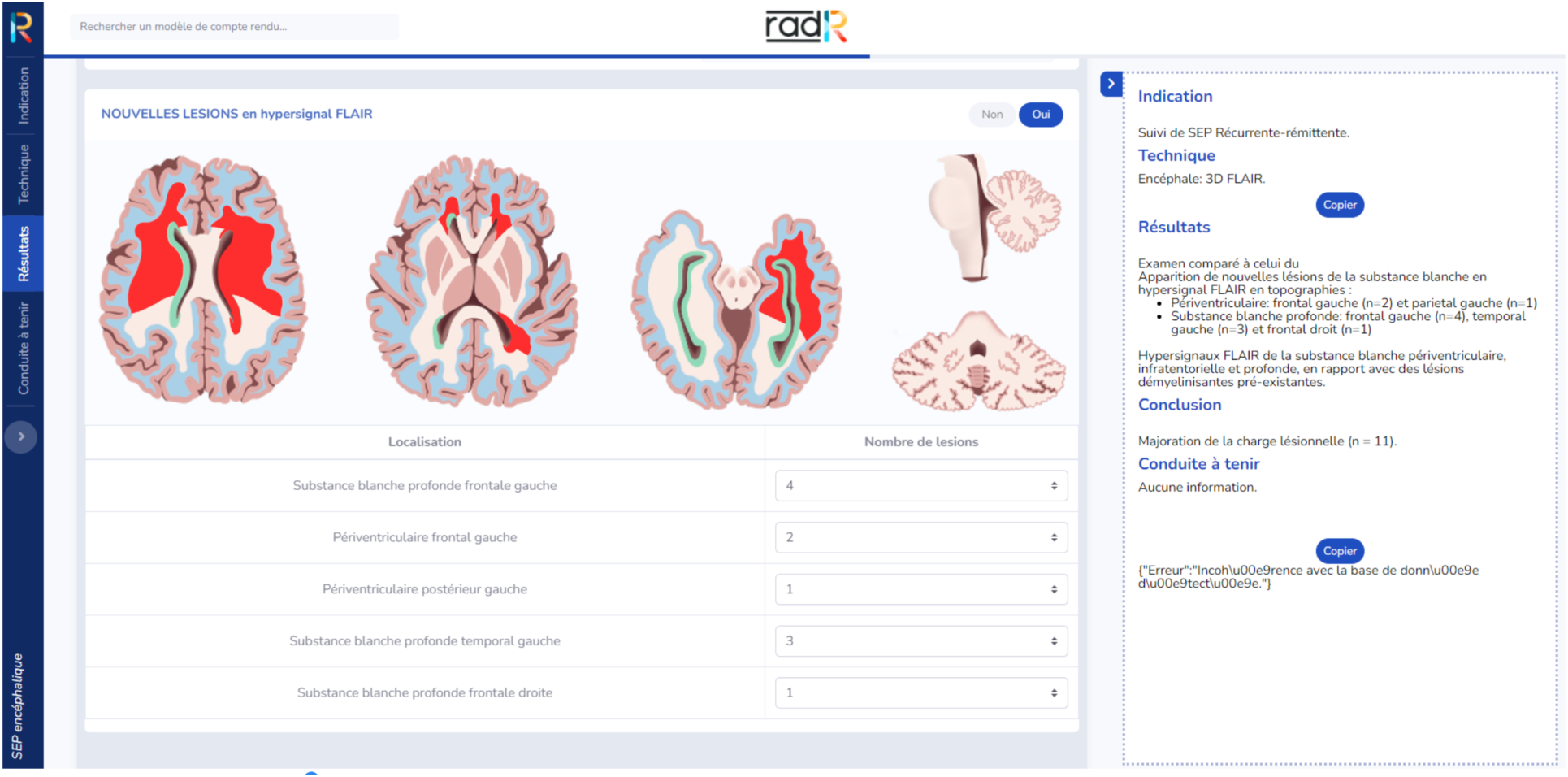
RadR Interface

### Non-structured report NSR

NSR will be performed as the radiologist own habits, using medical voice dication (Dragon)

### Variables

For the SR readings, the number of T2w-FLAIR hyperintensities, number of T1w enhancing lesions, localization of the lesions will be recorded through the exported .json file from RADR. Will be adjudicated by the two experienced raters that were not part of central readings : the presence of a new T2w hyperintensity, the presence of a T1w enhancement, the progression-activity of the disease status.

For the NSR, given the unconstrained design of reporting, chosen to approach a clinical time of reading/reporting, the counts of the lesions won’t be assessed. Only qualitative analysis will be made on the reports by the two experienced raters that were not part of central readings, extracting : the presence of a new T2w hyperintensity, the presence of a T1w enhancement, the progression-activity of the disease status. Each NSR will be stored according to the number of participation of the patient, to a separate informatics file.

For the CADR readings, failure or success of coregistration-fusion will be reported. The recorded times will be stored in a separate informatics file.

Clinical and demographical data will be extracted from patient informatics medical file and will be stored in a separate informatics file.

At the end of the readings all files will be made available to the two experienced neuroradiologist to adjudicate the data, and then provided to the statistician for the analysis.

### Primary endpoint

The main objective of this study is to evaluate the diagnostic performance of CADR compared to CR in the follow-up of MS.

### Secondary endpoints

The secondary objectives will be studied using a quantitative and qualitative analysis of the reading :

- to evaluate the diagnosis performance of readings process
- to evaluate the variability of readings
- to evaluate the variability of reportings
- to evaluate the impact of CADR on the variability of readings
- to compare the variability of readings according to the reader experience / radiologist/non-radiologist status
- to detail the variability of readings according to lesion localization using SR
- to evaluate the impact of CADR on the reading time
- to evaluate the impact of SR on the reporting time
- to evaluate the acceptability of CADR

### Primary judgment criteria

The main judgment criteria will be : Diagnostic accuracy (effectiveness) to assess the progression-activity of the disease based on the appearance of a T2w-FLAIR hyperintensity or a T1w enhancement on the follow-up examination, compared between consensus corrected CADR and CR readings.

### Secondary judgment criterion

The secondary criterion will be based on the count of T2w hyperintensities, count of T1w enhancement (quantitative), progression-activity status (qualitative):

- AUC-ROC, Sensitivity, Specificity, Positive Predictive Value, Negative Predictive Value
- Intra and Inter-observer agreements between CADR-SR and CR-SR / CADR-NSR and CR-NSR
- Intra and Inter-observer agreements between CADR-SR and CADR-NSR / CR-SR and CR-NSR
- Intra and Inter-observer agreements between CADR-SR and CR-SR
- Intra and Inter-observer agreements between CADR-SR and CR-SR in the subgroups of experiences
- Intra and Inter-observer agreements for lesion localisations using SR
- Reading times
- Reporting times
- Number of unexpected interruptions of the software, presence of a sub-optimal analysis described by the software.

## Statistics

### Number of subject needed

According to Zopfs et al. (10.) who retrospectively assessed 70 follow-up MRI of 53 patients with MS using the same CADR software, diagnostic accuracy increased from 67 to 90% for a and from 70 to 87% for the other using CADR versus CR. Diagnostic accuracy (effectiveness), is the a proportion of correctly classified subjects (True Positive TP +True Negative TN) among all subjects (TP+TN+False Positive FP+False Negative). According to the literature prevalence and contingency table (Zopfs et al. (10.)) of changes between follow-up exams, the expected effect of a 20% change of diagnostic accuracy, the number of subject needed to depict this difference with an absolute precision of 0.10 is beyond 63 and 95.

### Analysis

Statistics will be performed by an independent statistician, using STATA (MP 16, StataCorp). Quantitative variables will be presented as means and medians (interquartile range [IQR]), and categoric variables, as percentages. Mixed models will fit to compare the 4 reading methods (CADR-SR; CADR-NSR; CR-SR; CR-NSR) : A Poisson regression will be used for the number of new lesions (CADR-SR and CR-SR); a logistical regression, for binary variables such as the presence of at least 1 new lesion/state of progression-activity of the disease (all the readings); and a γ regression, for the reading time. The κ coefficient (qualitative) and intraclass correlation coefficient (quantitative) will be used to assess interobserver and intraobserver agreement and will be interpreted as suggested in previous publications (Landis and Koch): excellent agreement (κ ≥ 0.8), good agreement (κ ≥ 0.6), moderate agreement (κ ≥ 0.4), poor agreement (κ ≤ 0.4); and poor, fair, good, and excellent agreement categories will be qualified according to Cicchetti. The difference between dichotomous variables will be evaluated using McNemar’s test and the difference between continuous variables using the Wilcoxon test. Statistical significance will be set to p ≤ 0.05.

## Data Availability

no data available

## Notes

### Competing Interest Statement

The authors have declared no competing interest.

### Funding Statement

no funding

### Author Declarations

Comite d'Ethique pour la Recherche en Imagerie Medicale CERIM IRB Number: CRM-2206-271

